# Metabolomic signatures of lipid-modifying therapies using drug target Mendelian randomization

**DOI:** 10.1101/2021.08.06.21261699

**Authors:** Tom G Richardson, Genevieve M Leyden, Qin Wang, Joshua A Bell, Benjamin Elsworth, George Davey Smith, Michael V Holmes

## Abstract

**Background:** Large-scale molecular profiling and genotyping provide a unique opportunity to systematically compare the genetically predicted effects of therapeutic targets on the human metabolome.

**Methods:** We firstly constructed genetic risk scores for 8 drug targets on the basis that they primarily modify low-density lipoprotein (LDL) cholesterol (HMGCR, PCKS9 & NPC1L1), high-density lipoprotein (HDL) cholesterol (CETP), or triglycerides (APOC3, ANGPTL3, ANGPTL4 & LPL). We then used Mendelian randomization to evaluate the effect of each score on coronary artery disease (CAD) risk, and to systematically estimate their effects on 249 metabolic traits derived using blood samples from an unprecedented sample size of up to 115,082 UK Biobank participants.

**Results:** There was strong evidence of an effect of drug-based genetic scores on CAD risk with the exception of ANGPTL3. Genetically predicted effects on the blood metabolome were generally consistent amongst drug targets which were intended to modify the same lipoprotein lipid trait. For example, the linear fit for the MR estimates on all 249 metabolic traits for genetically-predicted inhibition of LDL cholesterol lowering targets HMGCR and PCSK9 was r^2^=0.91. In contrast, comparisons between drug classes that were designed to modify discrete lipoprotein traits typically had very different effects on metabolic signatures (e.g. HMGCR vs all 4 triglyceride targets had r^2^<0.02), despite largely consistent effects on risk of CAD. Furthermore, we highlight this discrepancy for specific metabolic traits, for example finding that LDL cholesterol lowering therapies typically had a weak effect on glycoprotein acetyls, a marker of inflammation (e.g. PCSK9: Beta=0.01, 95 CI%=-0.06 to 0.08, P=0.78). In contrast, all of the triglyceride modifying therapies assessed provided evidence of a strong effect on lowering levels of this inflammatory biomarker (e.g. LPL: Beta=-0.43, 95 CI%=-0.37 to -0.48, P=9×10^−50^).

**Conclusions:** Multiple lipid-modifying drug targets have therapeutically beneficial effects on CAD risk. Our findings indicate that genetically predicted perturbations of these drug targets on the blood metabolome can drastically differ, with potential implications for biomarkers in clinical development and measuring treatment response.

## Introduction

Cardiovascular disease (CVD), including coronary artery disease (CAD) and ischaemic stroke, is the leading cause of death worldwide^1^. Circulating lipoprotein lipid concentrations are of central importance to the aetiology of CAD^2, 3^. For example, clinical trials^4^ and studies of human genetics^5-7^ converge to support a causal role of apolipoprotein B (apoB) and low-density lipoprotein (LDL) cholesterol concentrations in the initial development and subsequent progression of CHD.

Pharmacological therapies that target the metabolism of blood lipids are routinely used for the prevention and treatment of CVD and are among the most widely prescribed medicines in the world ^8^. Interestingly, drug targets that modify concentrations of LDL cholesterol (e.g. statins, acting on HMG-CoA reductase [HMGCR]) and those designed to modify high-density lipoprotein (HDL) cholesterol (e.g. cholesteryl ester transfer protein [CETP] inhibitors) and triglycerides (e.g. angiopoietin like protein 3 [ANGPLT3] inhibitors) act on discrete pathways involved in lipid metabolism and thus while each drug class has proven^9-11^ or emerging^12-15^ efficacy for CVD risk reduction, their effects on the blood lipidome and metabolome are likely to vary considerably. This has implications, on understanding which biomarkers to measure (e.g. during clinical development in randomized controlled trials), and on gauging markers of treatment response^16^.

In this study, we sought to estimate the effects of lipid-modifying therapeutic targets on the blood metabolome to better characterize their impact on biomarkers related to CVD risk reduction. We constructed genetic instruments for drug targets that are either currently licensed or under development and grouped them according to their primary lipid of pharmacological focus: LDL cholesterol, HDL cholesterol, or triglycerides. We then compared the genetically predicted effects of therapeutic targets on CAD risk, before evaluating their effects on circulating lipoprotein lipid concentrations newly measured at large scale in the UK Biobank (UKB) study through the application of drug-target Mendelian randomization (MR)^17, 18^.

## Methods

### Instrument identification

Genetic instruments for each lipid modifying drug target were selected by undertaking genome-wide association studies (GWAS) of lipoprotein lipid traits measured using a conventional biochemistry assay in UKB^19^. These included HDL cholesterol (field 30760), LDL cholesterol (field 30780) and triglycerides (field 30870). Details on genotyping quality control, phasing, and imputation in UKB have been described previously^20^. GWAS were undertaken on UKB participants after excluding individuals of non-European descent (based on K-mean clustering of K=4) and standard exclusions, including withdrawn consent, mismatch between genetic and reported sex, and putative sex chromosome aneuploidy. Individuals who had measures of metabolic traits derived from a newly available nuclear magnetic resonance (NMR) platform in UKB were also excluded from these GWAS to avoid overlap with outcome samples (a potential source of bias in MR due to overfitting^21^). LDL cholesterol, HDL cholesterol and triglycerides were normalised using inverse rank-normalisation such that their mean was 0 and their standard deviation was 1. We used the BOLT-LMM (linear mixed model) software with adjustment for age, sex, fasting status and a binary variable denoting the genotyping chip used in individuals (the UKBB Axiom array or the UK BiLEVE array)^22^. BOLT-LMM uses a linear mixed effect model to account for the population structure within UKB which is why principal components were not included as covariates in the model. All analyses were conducted under UKB application #15825.

Final instrument selection for all 8 drug targets was based on results obtained from the GWAS of HDL cholesterol (to instrument *CETP*), LDL cholesterol (to instrument *PCSK9, HMGCR* and *NPC1L1*^4^) and triglycerides (to instrument *APOC3, ANGPTL3, ANGPTL4* and *LPL*^15, 23^). A selection criteria of genetic variants with P<1×10^−6^ which were located within a 100kbs region either side of encoding genes was applied to select instruments. We conducted linkage disequilibrium (LD) pruning for variants such that they had r^2^<0.1 using a reference panel of 503 European individuals enrolled in the 1,000 Genomes Project phase 3 (version 5)^24^. We additionally set out to identify genetic instruments for *PPARA* as a proxy for triglyceride modification through fibrates, although our GWAS only identified a single variant associated with triglyceride levels at this gene’s locus. We did not carry this target forward into downstream analyses given the challenges for genetic confounding of conducting gene-centric MR analyses with a single genetic instrument, particularly in gene dense regions where the function of each gene is not well understood, which may hinder inference^25^.

### Genome-wide association studies of metabolic traits and coronary artery disease

We applied the same GWAS pipeline described above to all 249 metabolic traits measured by targeted high-throughput NMR metabolomics from Nightingale Health Ltd (biomarker quantification version 2020) in UKB. These analyses were conducted under UKB project #15825. Measures were taken using non-fasting EDTA plasma samples (aliquot) obtained from a random subsample of 121,584 UKB participants. Sample sizes on the 249 metabolic traits for GWAS after QC ranged between n=110,051 to n=115,082 UKB participants. A full summary of sample sizes can be found in **Supplementary Table 1**. Each metabolic trait was normalised to have a mean of 0 and standard deviation of 1 using inverse rank-normalisation as above allowing comparisons to be made between derived effect estimates. As before, all GWAS were adjusted for age, sex, fasting time and genotyping chip.

Amongst these biomarkers were various lipoprotein lipids and their concentrations within 14 subclasses, fatty acids, ketone bodies, glycolysis metabolites and amino acids (see **Supplementary Table 1**). Further details have been described previously^26^. Ethical approval for this study was obtained from the Research Ethics Committee (REC; approval number: 11/NW/0382) and informed consent was collected from all participants enrolled in UKB.

Genome-wide summary-level estimates on coronary artery disease (CAD) risk were obtained from a previously conducted GWAS from the CARDIoGRAMplusC4D consortium^27^. CAD cases in this consortium were defined as myocardial infarction, acute coronary syndrome, chronic stable angina, or coronary stenosis >50%. In total there were 60,801 cases and 123,504 control assembled by CARDIoGRAMplusC4D across 48 studies which did not include the UK Biobank study.

### Statistical analysis

#### Drug-target Mendelian randomization

Univariable MR analyses were firstly undertaken to estimate the genetically predicted effects of each therapeutic target on risk on CAD (**Figure 1**). Estimates were derived by applying the inverse variance weighted (IVW) method whilst accounting for the correlation between instruments using the same reference panel as above^28, 29^. Further details on this approach have been described previously^28^. Briefly, we calculated the pairwise correlations between all variants included in genetic scores. These were then incorporated into the standard error terms of test statistics for the summary-level weighted generalized linear regression MR models.

**Figure 1.**
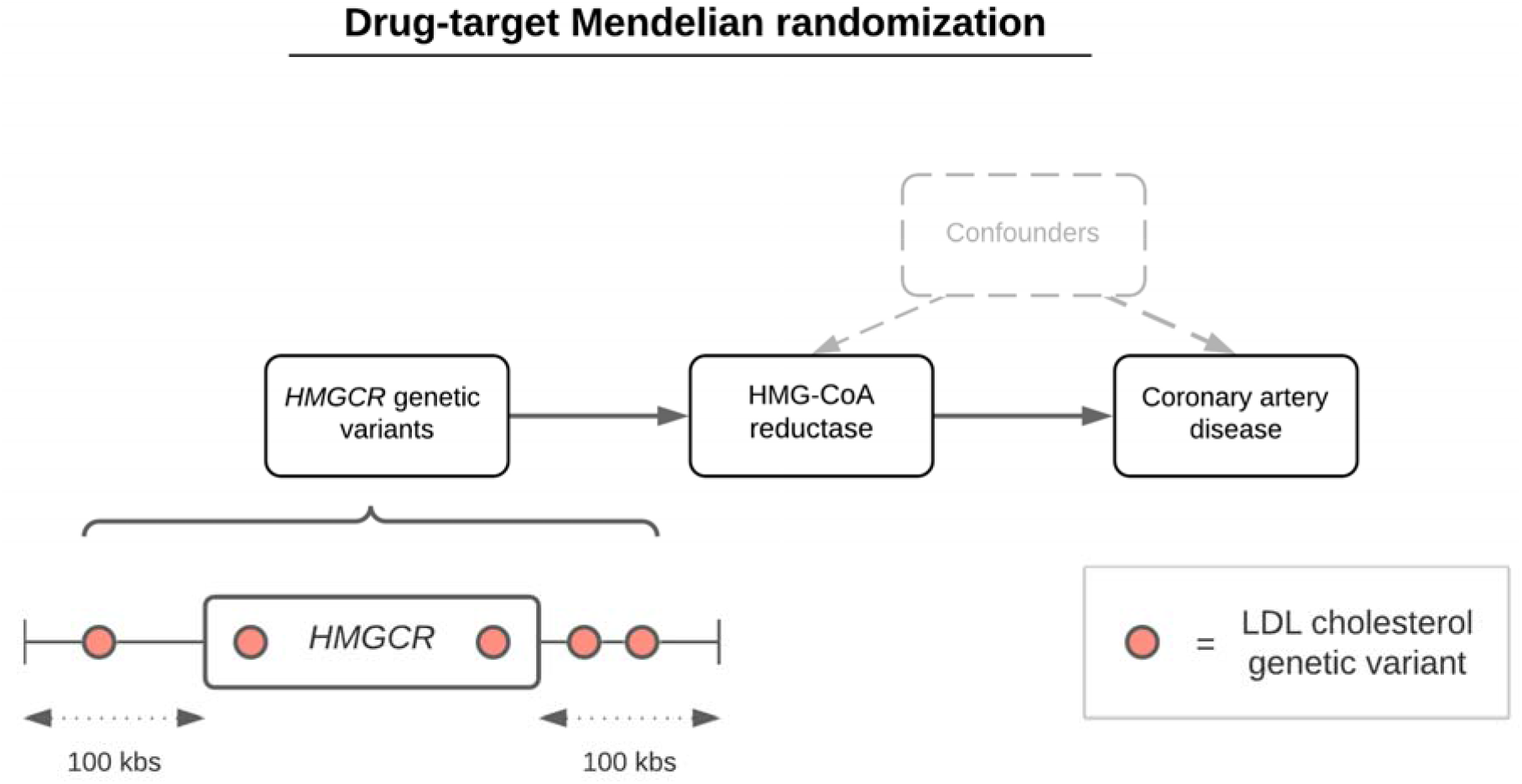
A schematic representation of the drug-target Mendelian randomization approach undertaken in this study using, e.g. HMGCR variants to proxy for HMG-CoA reductase inhibition (a proxy for statin therapy) to estimate its genetically predicted effect on coronary artery disease. Genetic variants robustly associated with a lipoprotein lipid trait (e.g. LDL cholesterol) based on P<1×10^−6^ within 100 kilobases (kbs) of encoding genes were identified as genetic proxies for perturbing therapeutic targets. A sensitivity analysis restricted to 50 kbs on either side of encoding genes was also undertaken in this study.

This approach was then applied systematically to estimate the genetically predicted effects of each target on each of the 249 metabolic traits in turn. Analyses were conducted in a two-sample data setting to ensure that our sample of UKB participants from which instruments were identified (i.e. the non-NMR subset of UKB) did not overlap with individuals analysed in the GWAS of metabolic traits in UKB. To account for multiple testing, false discovery rate (FDR) corrections were applied for each drug target analysed as a heuristic to highlight the most noteworthy findings given current sample sizes, although all results are reported in supplementary materials. We repeated this analysis restricting our instrument selection criteria to a 50kbs window around encoding genes for targets to assess the robustness of our findings to genetic confounding (i.e. variants influencing metabolic traits via neighbouring genes).

#### Comparison between different drug target effects across metabolome-wide traits

We initially compared effect estimates for CAD with a subset of lipid concentrations across drug targets using forest plots. More comprehensive comparisons on metabolome-wide results (i.e. on all 249 traits) were illustrated using scatter plots proposed previously to compare pairwise estimates between two targets with metabolic traits coloured based on their subcategories^30^. For comparative purposes, we scaled all metabolite estimates using a scaling factor based on each target’s corresponding genetically predicted effect on CAD risk. We used *HMGCR* estimates as our baseline comparison for each of the other 7 drug targets given the widespread adoption of statin therapy to treat individuals at elevated risk of cardiovascular disease. Comparisons between *HMGCR* estimates and those for each of the other scores were evaluated using generated R^2^ values as applied previously^30^. R^2^ values are the coefficients of determination estimating the quotient of the variances of the fitted values and observed values of the dependent variable using a linear regression model. Here, it describes the linear fit for the estimates on metabolic traits between the two drug targets assessed.

As a sensitivity analysis, we used individual-level data from UKB to investigate whether the genetically predicted effects of drug targets on lipoprotein lipid concentrations varied amongst participants subgroups stratified by their age. As described previously^31^, this approach permits the investigation of whether putative contingent factors in UKB may influence conclusions without directly conditioning of them. For example, in this study we might anticipate that the influence of statin medications on metabolic markers may distort effect estimates. However, adjusting for this factor either as a covariate or by stratifying participants on it is likely to induce collider bias into analyses which is recognised to potentially undermine causal inference^32^. As such we partitioned the unrelated European sample from UKB into the youngest (range from 40 to 54 years) and oldest (range from 61 to 71 years) subgroups (both n=30,000), where the number of reported participants taking statin medications was 5.6% and 27.7% respectively. Instruments for drug targets were then constructed as genetic risk scores using individual-level data from UKB and analysed against each measure of lipoprotein lipid concentrations in turn using linear regression adjusted for age, sex and the top 10 principal components.

All plots in this study were generated using the R package ‘ggplot2’^33^. MR analyses were conducted using the R package ‘MendelianRandomization’^34^. All analyses were undertaken using R (version 3.5.1).

## Results

### Genetic instrumentation of lipid modifying drug targets to estimate their therapeutic effects on coronary artery disease risk

GWAS of biochemistry measured LDL (n=328,111), HDL (n=300,528) and triglyceride (n=328,498) in UKB identified a total of 137 weakly independent instruments (at r^2^<0.1 across the 8 lipid modifying targets evaluated in this study (**Supplementary Table 2**). Each target provided strong evidence of a genetically predicted effect on CAD risk (based on FDR<5%) with the exception of *ANGPTL3* (**Supplementary Table 3** and **Figure 2**), in keeping with prior findings^5, 30, 35-38^. F-statistics did not indicate that drug target scores were prone to weak instrument bias (F=58.3 to 297.1) (**Supplementary Table 3**).

**Figure 2.**
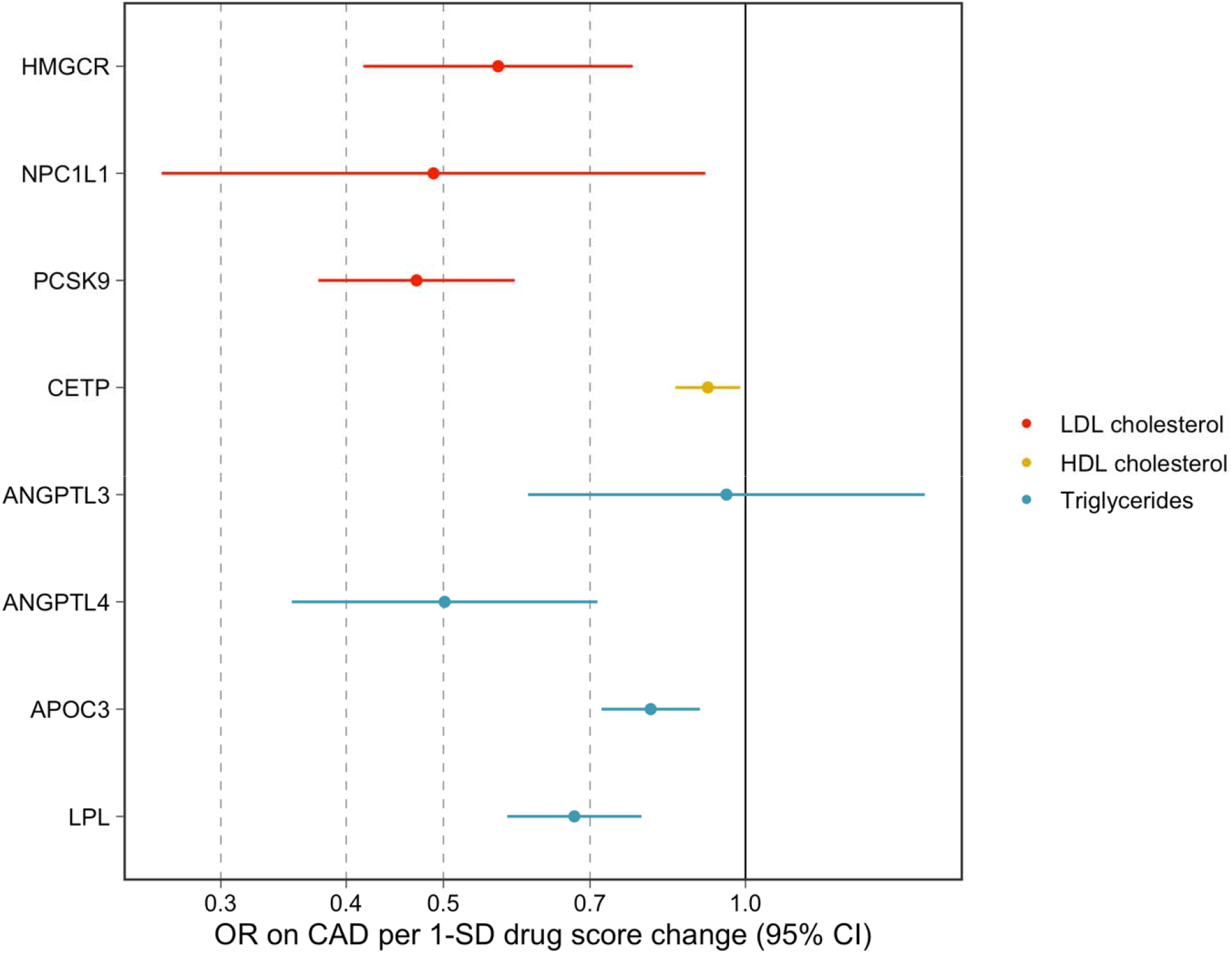
A forest plot visualizing the genetically predicted effects of lipid modifying drug targets on risk of coronary artery disease (CAD) using Mendelian randomization. Estimates are colour coded based on the lipoprotein lipid trait estimates used to derive genetic scores. Each genetic score was oriented to mimic the putative effects of drug targets on lipoprotein traits, meaning that effect estimates correspond to odds of CAD per 1 standard deviation (SD) change in either lower LDL cholesterol, higher HDL cholesterol or lower triglyceride levels via each specific drug target. Note that in the case of CETP, we are not ascribing causal effects to HDL cholesterol – rather, we are orientating CAD effect estimates corresponding to a genetically-predicted increase in HDL cholesterol arising from pharmacological inhibition of CETP.

### Systematic evaluation of genetically predicted therapeutic target effects on metabolic traits

In total, there were 2,814 genetic variants robustly associated with at least one measure (based on P<5×10^−8^) across 721 independent genetic loci (as reported previously) (**Supplementary Table 4**). All of the 249 metabolic traits were represented amongst these findings (i.e. each trait quantified by the NMR platform had at least one SNP association at GWAS levels of significance) with the majority having dozens of independent variants associated with their levels (median: 74 variants, interquartile range: 27 variants) (**Supplementary Table 5**). Systematically estimating genetically predicted effects of each lipid modifying target in turn on each of the 249 metabolic traits identified a total of 1,588 effects robust to FDR<5% corrections (**Supplementary Tables 6 to 13**). Investigating the robustness of our results to a more stringent instrument selection criteria (i.e. 50kbs either side of encoding genes as compared to 100kbs in our main analyses) provided strong evidence of homogeneity between genetically predicted drug-target effects in the original analysis (**Supplementary Figures 1 to 8**).

A subset of these estimates related to lipoprotein particle, cholesterol and triglyceride concentrations across the 8 drug targets have been highlighted in **Figure 3**. Broadly, the LDL cholesterol modifying targets (*HMGCR, PCSK9* and *NPC1L1*) provided evidence of genetically predicted effects on lower levels of very low-density lipoprotein (VLDL), intermediate density lipoprotein (IDL) and LDL related particle concentrations, but typically weak evidence on HDL related markers (with the exception of very large HDL particles, for which genetics instruments for both HMGCR and PCSK9 showed strong evidence of lowering). For example, the strongest evidence identified using the PCSK9 score was on large LDL particle concentrations (Beta=0.96 SD reduced per 1-SD lowering in LDL cholesterol, 95% CI=0.87 to 1.04, P=3×10^113^). The concentration of cholesterol within lipoprotein particle subclasses tended to mimic the associations identified for lipoprotein particle concentration. In contrast, generally weaker effects of genetic instruments for HMGCR, PCSK9 and NPC1L1 were identified for triglyceride concentrations within the same lipoprotein particle subclasses.

**Figure 3.**
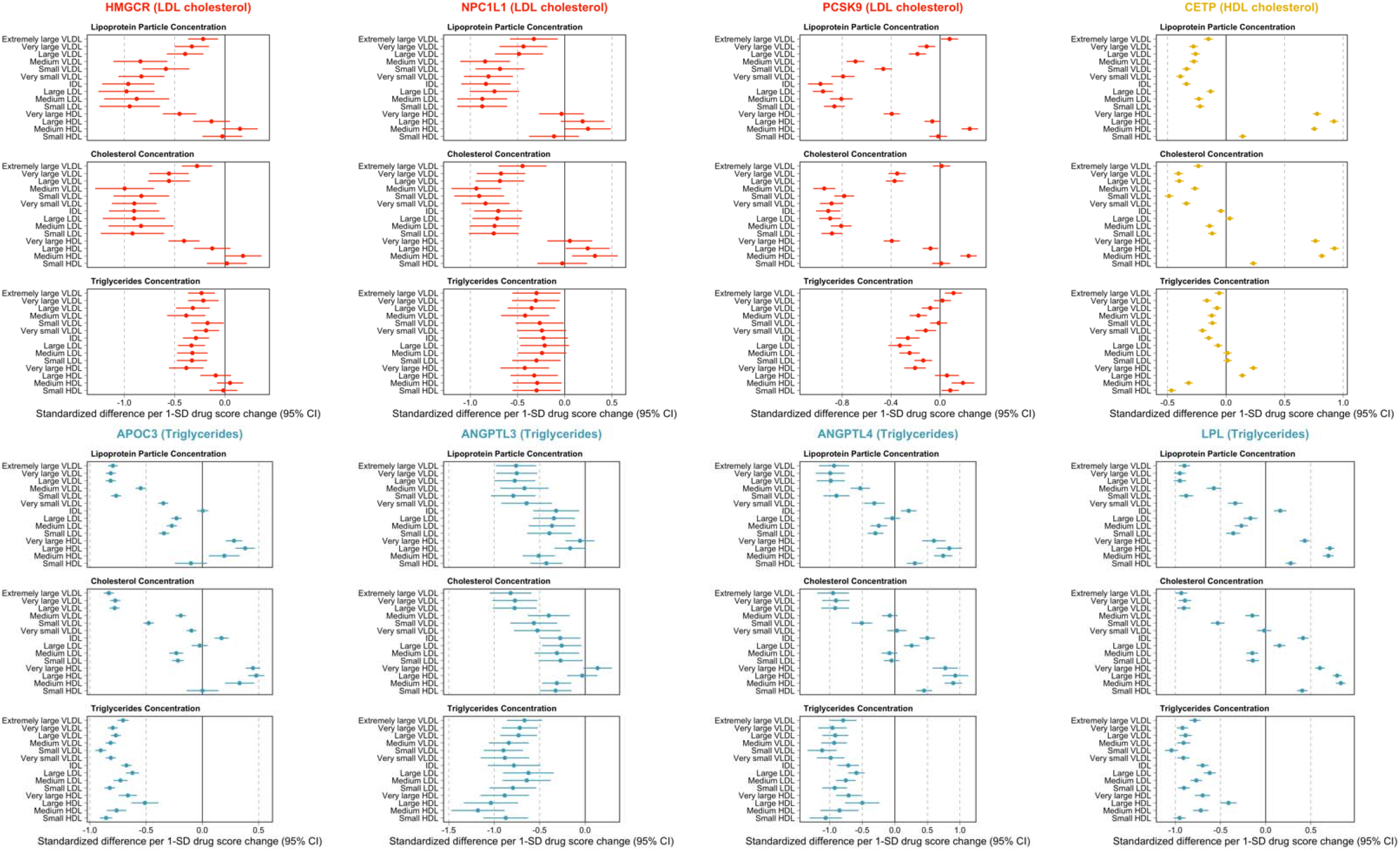
Forest plots illustrating the genetically predicted effects of lipid modifying drug targets on measures of circulating metabolite concentrations using nuclear magnetic resonance in the UK Biobank study. Effect estimates are based on a standard deviation (SD) change in the genetically predicted drug target scores oriented to reflect therapeutic intervention (i.e. lower LDL cholesterol, lower triglycerides and higher HDL cholesterol). Scores were derived using genetic variants robustly associated which lipoprotein lipid traits (as indicated in each target’s legend) at each encoding gene’s region.

Orientated to a lowering of CHD risk, the HDL cholesterol modifying target *CETP* provided evidence of lower genetically predicted effects on VLDL and LDL circulating metabolite concentrations. Notably, comparatively larger effects were identified on lipoprotein particle concentration and cholesterol concentrations within HDL subclasses with positive associations identified for these HDL-related traits. In contrast, marked heterogeneity was found in relation to triglycerides concentrations, with genetically predicted estimates suggesting an effect on higher very large and large HDL-C concentrations and on lower levels of medium and small HDL concentrations.

Genetically predicted triglyceride modifying targets (*APOC3, ANGPTL3, ANGPTL4* and *LPL*) markedly lowered triglyceride concentrations across the spectrum of lipoprotein subclasses – this was in contrast to genetics instruments for HMGCR, PCSK9, NPC1L1 and CETP where effect estimates were weaker and tended to be on both sides of the null). For lipoprotein particle and cholesterol concentrations, lowering effects of triglyceride modifying targets were typically found for the larger metabolic traits (i.e. VLDL), with an inflection point at IDL seen for *ANGTPL4, APOC3* and *LPL* but not for ANGPTL3.

A comparison of these analyses repeated in the youngest and oldest subgroups using individual-level data from unrelated individuals within UKB (both n=30,000) can be found in **Supplementary Figure 9**. Overall metabolic signatures did not appear to drastically differ between these strata defined by age, suggesting that treatment with statins was unlikely to lead to major perturbations in the effect estimates we present. Whilst overall trends did not typically vary from those identified in the full sample, these findings suggest that analyses which directly adjust for contingent factors within UKB, such as statin medications, are likely to introduce collider bias into their findings (as proposed previously^31^).

We also identified differing signatures between drug target classes for non-lipoprotein lipid related traits. For instance, LDL lowering targets typically provided weak evidence of a genetically predicted effect on glycoprotein acetyls (GlycA), a marker of inflammation (e.g. PCSK9: Beta=0.01, 95 CI%=-0.06 to 0.08, P=0.78). In contrast, all triglyceride lowering targets provided strong evidence of a genetically predicted effect on lowering GlycA (e.g. LPL: Beta=-0.43, 95 CI%=-0.37 to -0.48, P=9×10^−50^). Full results from this systematic evaluation on each of the 249 metabolic traits can be found in Supplementary Tables 6 to 13.

### Genetically predicted metabolic effects for drug targets in comparison to statin medication

We systematically compared the genetically predicted effects of each drug target on all 249 metabolic traits with HMGCR acting as a proxy for statin therapy. For comparative purposes, estimates were scaled in accordance with their respective effect estimates on CAD as reported in **Supplementary Table 3**. In general, we identified strong evidence of concordance between the other LDL cholesterol lowering therapies (*PCKS9* and *NPC1L1*) with the *HMGCR* score (r^2^=0.91 & r^2^=0.79 respectively). **Figure 4a** illustrates the linear trend identified between genetically predicted effects of PCSK9 and HMGCR on metabolic markers. **Supplementary Figure 10** contains the corresponding plot for *NPC1L1*.

**Figure 4.**
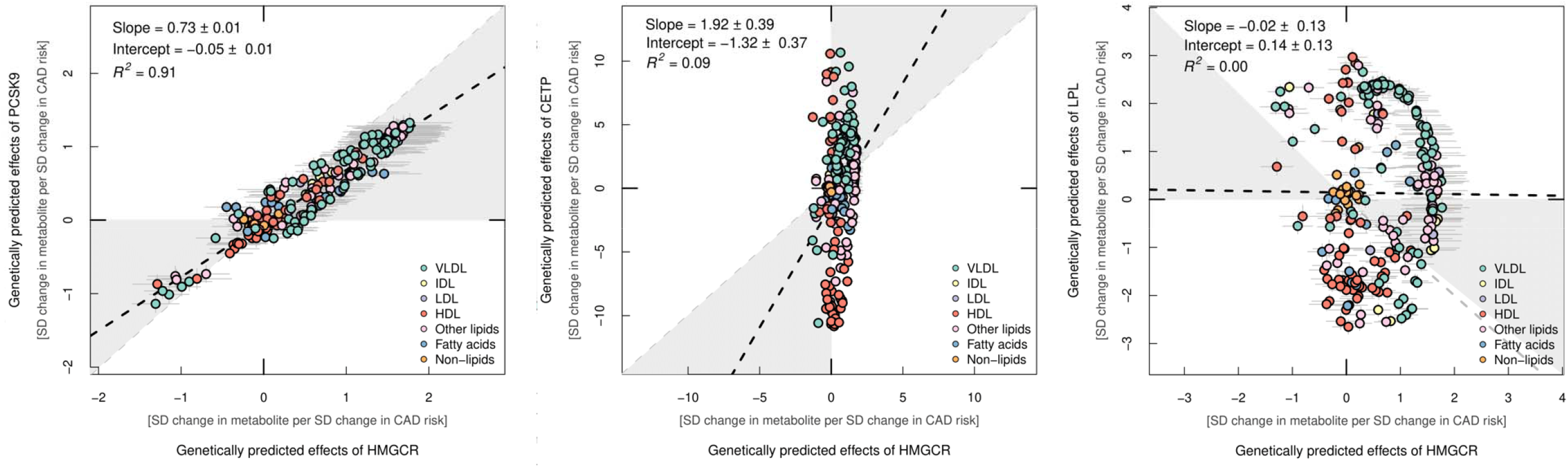
A comparison of distributions between genetically predicted drug target effects A) PCSK9 B) CETP and C) LPL on metabolic traits using nuclear magnetic resonance in the UK Biobank study. In each figure estimates are compared with the results from the HMGCR score. Effect estimates were scaled in accordance with the corresponding effects of these genetically predicted drugs targets on risk of coronary artery disease which is why axes vary between plots. Points are coloured based on subcategories of metabolic traits indicated in the figure legends.

In contrast, comparisons with the HDL cholesterol raising (*CETP*) (r^2^=0.09) (Figure 4b) and triglyceride lowering (*APOC3, ANGPTL3, ANGPTL4* and *LPL*) (all r^2^≤0.02) targets provided weak evidence of concordance with the *HMGCR* score. Figure 4c visualises this general lack of concordance using the *LPL* and *HMGCR* score comparison as an exemplar. Broadly, both the *LPL* and *HMGCR* scores provided evidence of genetically predicted effects on higher levels of various triglyceride-rich VLDL related traits (highlighted in green). However, conversely, the *LPL* score typically provided stronger evidence of an effect on HDL related traits (highlighted in red) which was generally not the case for the *HMGCR* score. As expected, both scores provided weak evidence of an effect on non-lipid related traits (highlighted in orange). All other figures generated from this analysis on the other mentioned targets can be found in **Supplementary Figures 11 to 13**.

## Discussion

In this study, we explored the genetically predicted metabolic effects of modifying LDL, HDL, and triglycerides via drug targets that are either well-established, recently licensed and/or under development^39-42^. Our findings demonstrate that drug targets that principally act to modify LDL cholesterol (e.g. statins, PCSK9 inhibitors and ezetimibe) have broadly similar effects on the blood metabolome. In contrast, effects of drugs designed to modify HDL cholesterol and triglycerides had very different effects on metabolic biomarkers, even when scaled to the same difference in risk of CAD. These findings provide a catalogue of genetically-predicted pharmacological effects on the blood metabolome, which serves to illustrate the heterogeneity between different lipid modifying therapies, highlighting the need for rich phenotyping of lipoprotein lipids in developing assays that gauge treatment response.

Our findings illustrate the tapestry of metabolic biomarker associations that are predicted to be downstream consequences of pharmacological modification of a therapeutic target. While these findings do not provide evidence of causation of these metabolic biomarkers, rather, they employ drug target Mendelian randomization as a means of characterizing therapeutic effects on the metabolome^43, 44^. Such diverse effects can then potentially be triangulated^45^ to explore patterns of metabolomics where signatures are consistent with cardiovascular risk reduction. In-depth investigations into the independent causal role of specific metabolic traits at a granular level can then be explored using approaches such as multivariable MR^46^. For example, one might construct genetic instruments for biomarkers that are downstream consequences of HMGCR inhibition and conduct de novo multivariable MR analyses of these traits in order to identify the mediating mechanisms beyond apoB or LDL cholesterol. It may thus be possible to identify in finer detail which metabolic biomarkers are causally implicated in CVD and identification of such may be the focus of new therapeutic targets for clinical development. While previous studies, including those that we conducted, identified apoB as the fundamental driver of lipid-mediated CVD^7^, a greater understanding of the causal components should facilitate new avenues of investigation and resultant pharmacological development.

One of the striking findings is the general consistency of associations between drug targets and particle concentration and cholesterol concentration (likely owing to the high correlation between these phenotypic traits) and the divergence between particle and cholesterol concentration and triglycerides concentration. This was most notable when comparing drugs across their primary lipid indication – i.e. drugs that were developed on the basis of LDL cholesterol lowering tended to have modest associations with a general reduction in triglyceride concentrations across lipoprotein particles. In contrast, HDL cholesterol raising variants in *CETP* were identified to generally lower triglycerides in apoB containing lipoproteins whereas for HDL particles triglycerides was increased in very large and large HDL particles and reduced in medium and small HDL particles. For the drug targets where triglycerides metabolism was the primary lipid of pharmacological focus for development, triglycerides concentrations were lower across the lipoprotein particle spectrum. Since most of these drug targets demonstrated genetic evidence of CAD lowering, one might draw conclusions from such heterogeneity of triglycerides effects indicative that perhaps triglycerides was not important per se but rather the trait of interest was cholesterol or lipoprotein particle concentration (indexed e.g. by apoB concentrations). However, previous multivariable MR analyses that included triglycerides, apoB and LDL-C in the model demonstrated a direct effect of triglycerides consistent with a potential causal role of triglycerides in CAD^7, 47^ using the same dataset from the CARDIoGRAMplusC4D consortium as analysed in this study^27^. Thus, while our findings illustrate pronounced heterogeneity in cholesterol and triglyceride lipoprotein lipid concentrations arising from genetically-predicted pharmacological inhibition of lipid modifying drug targets, drawing causal conclusions from such perturbations is non-trivial and requires MR of the individual phenotypes, as described previously.

The findings presented here have been made available by large-scale phenotyping using NMR targeted metabolomics in UKB in combination with GWAS genotyping. Such data provide resolution of lipoprotein lipids at scale and enable genetic analyses of the type we present. The value of metabolomics may be to offer signatures of treatment response which can then be used to guide pharmacological treatment. Such may be of utility from an early stage – e.g. during phase I, II and III clinical trials, where biomarkers are often used as a means of measuring treatment response across different concentrations of drugs^17^, and post-marketing, when assessing interindividual response to treatment. Equally, our study has noteworthy limitations. For example, although previous studies have used similar criteria for instrument selection for the gene-based drug scores used in this study, we are unable to rule out genetic confounding as a potential source of bias in our analyses. Furthermore, we have used common genetic variants associated with lipoprotein lipid traits as a source of genetic instruments in this work. Future endeavors harnessing genetic effects on molecular traits (e.g. circulating proteins) or rare (and potentially highly penetrant) genetic variants may yield alternate strands of evidence to complement (or contradict) our results. In particular, these alternative approaches to genetic instrument selection for may identify a more valid proxy for targets such as ANGPTL3^48^.

In summary, our study characterises the repertoire of genetically-predicted lipid-modifying therapies on the blood metabolome. These findings demonstrate the widespread metabolic perturbance that arises from genetically-evaluated modifications of therapeutic targets and heterogeneity between discrete classes of drugs, especially when their primary lipid trait differs. Such findings may be useful to illustrate the utility of drug target Mendelian randomization in gauging the predicted effects of drugs on omics traits to guide dose-ranging studies during clinical development and as a marker of treatment response.

## Data Availability

All data analysed in this study is either available from the referenced public repositories or accessible via an approved application to the UK Biobank study.

https://www.ukbiobank.ac.uk/enable-your-research/apply-for-access

## Acknowledgements

We would like to thank the CARDIoGRAMplusC4D consortium for making their summary statistics available for the benefit of this work and to the participants of the UK Biobank study. Additionally, the authors are grateful to UK Biobank for access to data to undertake this study (Project #15825 and #30418). MVH works in a unit that receives funding from the UK Medical Research Council and is supported by a British Heart Foundation Intermediate Clinical Research Fellowship (FS/18/23/33512) and the National Institute for Health Research Oxford Biomedical Research Centre. George Davey Smith and Joshua Bell work in the Medical Research Council Integrative Epidemiology Unit at the University of Bristol (MC_UU_00011/1).

